# Enrollees and Early Lessons from an Agentic Healthcare Platform with Physicians in the Loop

**DOI:** 10.64898/2026.11.12.26367291

**Authors:** Zili Shen, Anu Ramachandran, Elaine Hsiang, Jasleen Gill, Ryan Miller, Nicholas Stark

## Abstract

Agentic healthcare platforms that combine large language models with physician oversight are growing quickly, but little is known about who uses them. We characterized 3,223 enrollees of Lotus AI, a free, physician-supervised agentic platform, who completed onboarding, messaged the agent, and had a linked health record between February and July 2026, and compared them with national benchmarks. Median age was 51 years and 62% were male. Chronic condition prevalence exceeded 2024 National Health Interview Survey estimates in every age band by 4 to 7 percentage points; 18% had a functional or symptom-defined disorder; among the 30% with four or more ambulatory visits in the prior year, care was spread across nearly five providers, with 44% meeting a threshold for high fragmentation. Users with functional disorders generated 1.8 times as many physician escalations and 2.6 times as many laboratory orders as other users, but no more prescriptions. This profile differs from the younger, predominantly female populations reported for direct-to-consumer telemedicine. It moves chronic disease management, functional disorder pathways, and care coordination from aspirations to design requirements, and we argue that deployed AI care platforms should characterize their populations against national benchmarks early.

## Introduction

Primary care faces growing challenges in meeting patient demand, maintaining continuity, and coordinating increasingly complex care. One estimate suggests that primary care physicians would require twenty-seven hours per day to deliver all care to their assigned patients, while the proportion of Americans with an established source of primary care has declined in the past two decades.^2,3^ Agentic healthcare platforms integrating large language models (LLMs) offer potential solutions to these challenges by reducing administrative burden, expanding access to care, and supporting guideline-concordant clinical decision-making.^4–8^ Approximately one in three US adults now use artificial intelligence (AI) chatbots for health-related information.^9^

Despite this rapid expansion, little is known about who receives care from agentic healthcare platforms. Existing evidence comes largely from clinician-delivered telemedicine and is conflicting. Studies of direct-to-consumer telemedicine have generally identified younger, healthier patients with fewer comorbidities,^10–12^ while other investigations have reported greater utilization among individuals with chronic conditions.^13^ These findings may not transfer to agentic healthcare platforms. Telemedicine still requires a clinician at every encounter, whereas agentic healthcare platforms leverage LLMs to offer continuous conversations on demand, including when traditional care is least available.^14^ Platforms that eliminate traditional barriers related to cost, insurance coverage, and appointment availability may serve substantially different populations.^9,15,16^ Regulatory reporting does not fill this gap: despite FDA draft guidance^17^, public summaries of FDA-authorized AI devices rarely report patient characteristics.^18,19^

Agentic healthcare platforms may, like direct-to-consumer telemedicine, attract younger and healthier users seeking convenience. Alternatively, they may disproportionately attract patients with multiple comorbidities, functional disorders, and fragmented care. In addition to managing complex health conditions, these patients must coordinate care across many clinicians^20^, and their health records are often scattered among multiple health systems.^21^ They also account for more primary care visit time and send more patient portal messages.^22,23^ Continuous asynchronous access and consolidated health data may be especially valuable to them, but this group may also be more vulnerable to triage and escalation errors.^24^ Because AI performance depends on the population in which it is deployed and LLM evaluations rarely use data from real patients,^25,26^ understanding the users served by agentic healthcare platforms is essential to enabling the rigorous evaluation of the safety, quality, and value of care provided.^18,27^

To address the relative lack of agentic healthcare platform consumer data, we conducted a retrospective case study to characterize enrollees of Lotus AI, an agentic healthcare platform that integrates LLMs to provide around-the-clock personalized health guidance, longitudinal patient engagement, and clinical triage with physician oversight and escalation when indicated. We categorized patients by age, sex, chronic disease burden, functional disorder burden, and care fragmentation, and compared our patient population with national benchmarks.

## The Platform

Lotus AI consists of two LLM agent loops (Figure 1). The patient-facing Lotus agent converses directly with users around the clock. At enrollment, patients consent to retrieval of their health record from a health information exchange (HIE) and patient portals; the agent then reasons over the linked record, synced wearable data, chat history, Lotus care history, and a curated knowledge base of clinical guidelines and literature. An escalation algorithm determines which requests require physician review. Escalations route to Cortex, a clinical decision support agent used by Lotus physicians with access to the same patient records. Escalations have three outcomes: lab orders, prescriptions, and follow-up messages.

**Figure 1.**
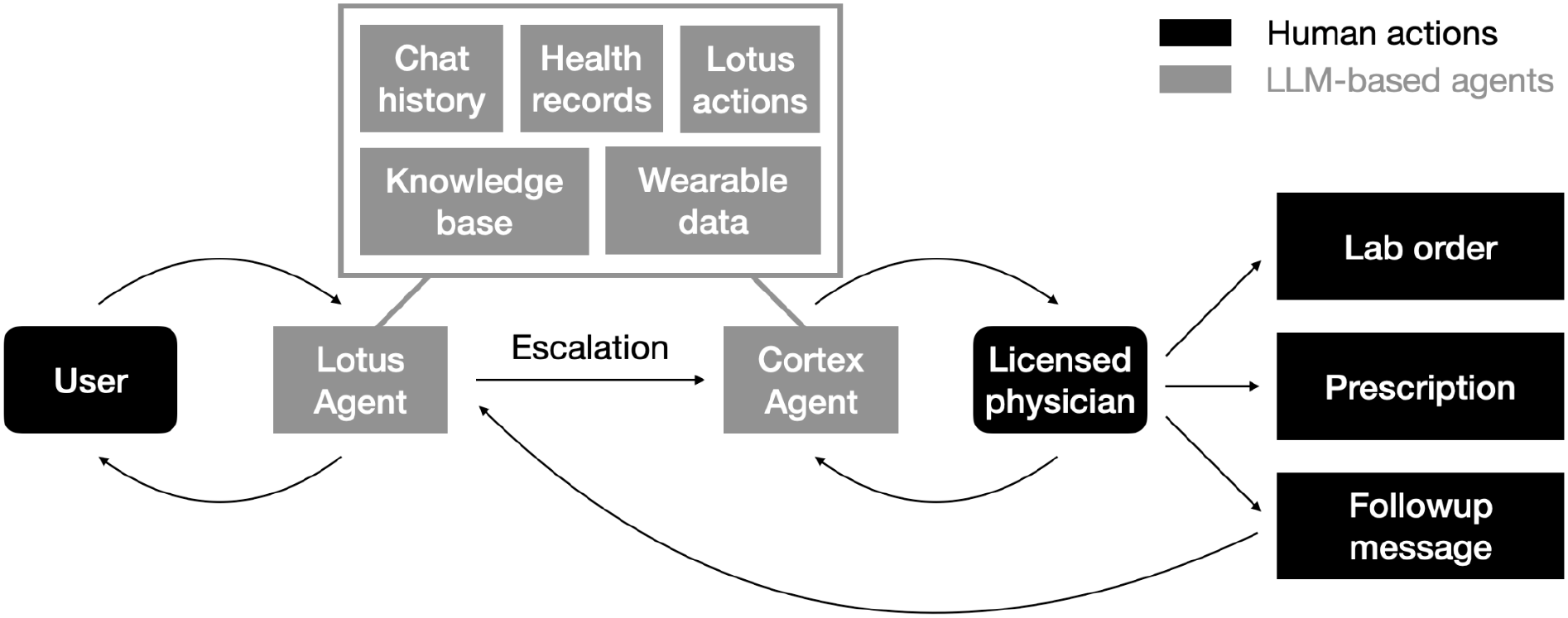
Lotus app system architecture. From left to right: users chat with Lotus Agent, an LLM with tools and access to user-specific chat history, health records, Lotus actions, wearable data, and a general knowledge base containing research papers and clinical guidelines. An escalation algorithm determines when to create a request for the Cortex Agent, which has access to the same data and can chat with licensed physicians. Physicians can create lab orders, prescription, and follow-up messages on the platform.

Cortex may suggest the best action but physician approval is always required. The service was free to patients during the study period.

## Methods

### Study Population

We included users who completed onboarding between February 1 and July 31, 2026, sent at least one message to the agent in that period, and had at least one ICD-10–coded condition dated on or after October 1, 2015 (the US ICD-10 transition), indicating both active use and successful record linkage. A condition’s date was its recorded date or, if absent, its onset date.

### Data collection

Date of birth and sex assigned at birth were collected from identity verification platforms and HIE. Age was calculated as 2026 minus birth year. We extracted Fast Healthcare Interoperability Resources Conditions, Encounters, and Practitioners for all cohort users from HIE and measured platform activity as the number of escalations, laboratory orders, and prescriptions during the study period. Advarra IRB reviewed the study (Pro00094483) and determined it exempt.

### Chronic and Functional Conditions

We mapped the 10 chronic conditions in the 2024 National Health Interview Survey (NHIS) to ICD-10 codes (Table S1) and applied them to Condition records with usable dates and ICD-10 coding. We defined fourteen functional somatic disorders from the literature: irritable bowel syndrome, fibromyalgia, myalgic encephalomyelitis/chronic fatigue syndrome, functional dyspepsia, functional constipation, functional diarrhea, cyclic vomiting syndrome, interstitial cystitis/bladder pain syndrome, functional neurologic disorder, postural orthostatic tachycardia syndrome, mast cell activation syndrome, long COVID, temporomandibular disorder, and complex regional pain syndrome type 1 (Table S2).

### Care Fragmentation analysis

For the year before the study period (February 1, 2025, to January 31, 2026), we identified ambulatory evaluation and management visits (CPT 99202–99215) with an associated provider’s National Provider Identifier (NPI). Following established guidance, we assessed fragmentation only among users with at least four such visits,^28^ calculating the usual provider of care (UPC) index^29^ (share of visits to the most frequently seen provider) and the reversed Bice–Boxerman continuity of care index (rBBI = 1 − BBI),^30^ the probability that two randomly selected visits were with different providers. We classified rBBI above 0.85 as high, 0.70 to 0.85 as medium, and below 0.70 as low fragmentation.^31^

### Statistical Analysis

Age-band prevalences are reported with Wilson 95% confidence intervals, and differences from published NHIS estimates with MOVER 95% confidence intervals,^32,33^ which combine our Wilson intervals with the NHIS design-based intervals. Binary outcomes were compared with Fisher’s exact test and summarized as conditional maximum-likelihood odds ratios with exact 95% confidence intervals. Adjusted odds ratios came from logistic regression (Wald 95% confidence intervals), adjusting for age and sex and, for high fragmentation, the logarithm of the number of provider visits, since rBBI depends on visit count. Count outcomes were compared with the two-sided Mann–Whitney U test, and 95% confidence intervals for ratios of means and for each group’s share of total activity were estimated by a nonparametric bootstrap with 2,000 user-level resamples and the percentile method. All p values are two-sided; analyses were performed in Python with SciPy and statsmodels.

## Results

### Study population

A total of 3,223 users met inclusion criteria. Median age was 51 years, and 62.1% were male (Table 1). These users had 12,219 eligible visits (83.4% had an associated NPI) and 430,442 eligible Condition records.

**Table 1.** Lotus user demographic distribution.

| **Characteristic** | **n** | **%** |
|---|---|---|
| **Total population** | **3,223** | **100.0** |
| **Age, years** |  |  |
| 18–24 | 158 | 4.9 |
| 25–34 | 484 | 15.0 |
| 35–44 | 577 | 17.9 |
| 45–54 | 613 | 19.0 |
| 55–64 | 650 | 20.2 |
| 65–74 | 466 | 14.5 |
| 75+ | 275 | 8.5 |
| Missing | 0 | 0.0 |
| **Sex** |  |  |
| Female | 1,219 | 37.8 |
| Male | 2,001 | 62.1 |
| Missing | 3 | 0.1 |

### Chronic and functional disorder prevalence

Overall, 1,906 users (59.1%) had at least one NHIS chronic condition and 584 (18.1%) had a functional disorder. In every age band, chronic condition prevalence exceeded 2024 NHIS estimates (Figure 2): 33.2% vs. 26.2% at 18 to 44 years (difference, 7.0 percentage points; 95% CI, 4.2 to 9.9), 65.6% vs. 61.5% at 45 to 64 years (4.1; 1.2 to 6.9), and 90.8% vs. 86.5% at 65 years or older (4.3; 1.9 to 6.4).

**Figure 2.**
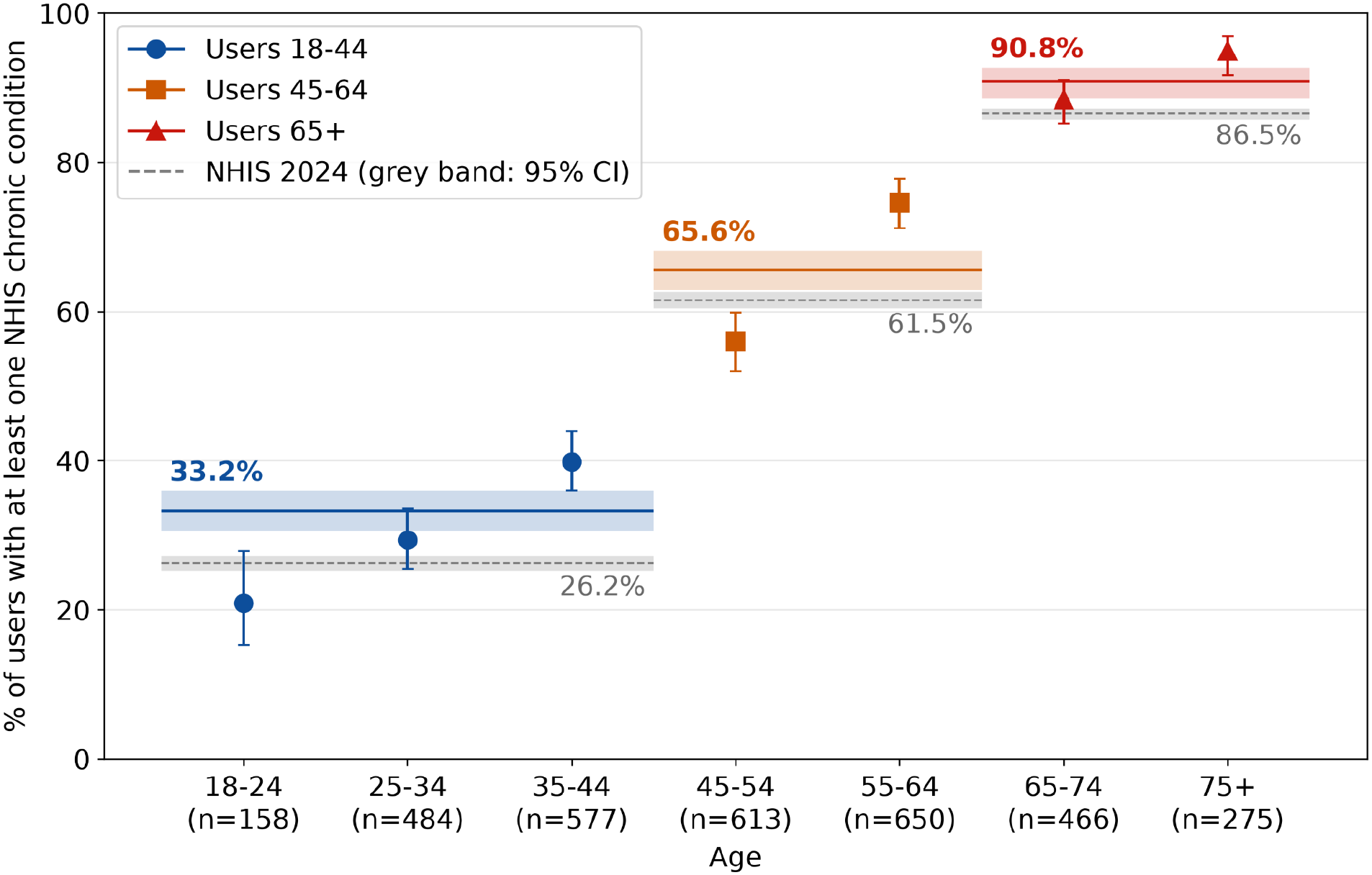
Chronic condition prevalence among Lotus users, stratified by age. The figure shows the percentage of users with at least one chronic condition, as defined by the National Health Interview Survey (NHIS), by age group. Markers show the percentage in each age group, with error bars indicating 95% confidence intervals; the number of users in each age group is shown below its label. Colored horizontal lines show the percentage for users aged 18 to 44, 45 to 64, and 65 years or older, with shaded bands indicating 95% confidence intervals and the value labeled above each band. Dashed gray lines show the corresponding 2024 NHIS national estimates for the same age bands, with gray bands indicating 95% confidence intervals and the value labeled below each band.

### Platform activity among users with functional disorders

Users with a functional disorder generated more escalations (mean, 2.62 vs. 1.47 per user; ratio of means, 1.78; 95% CI, 1.42 to 2.21; P<0.001) and were more likely to have any escalation (47.4% vs. 41.7%; OR, 1.26; 95% CI, 1.05 to 1.52; P=0.01). The excess was in testing, not prescribing (Figure 3). Laboratory orders were placed for 11.1% vs. 6.6% (OR, 1.77; 95% CI, 1.29 to 2.41; P<0.001), at 2.6 times the per-user rate (0.96 vs. 0.37; 95% CI for ratio, 1.67 to 3.93); users with a functional disorder were 18.1% of the cohort but accounted for 36.6% (95% CI, 26.3 to 46.6) of all laboratory orders. Prescriptions did not differ (9.8% vs. 11.1%; OR, 0.86; 95% CI, 0.63 to 1.17; P=0.38; 0.48 vs. 0.50 per user; ratio, 0.97; 95% CI, 0.48 to 1.68).

**Figure 3.**
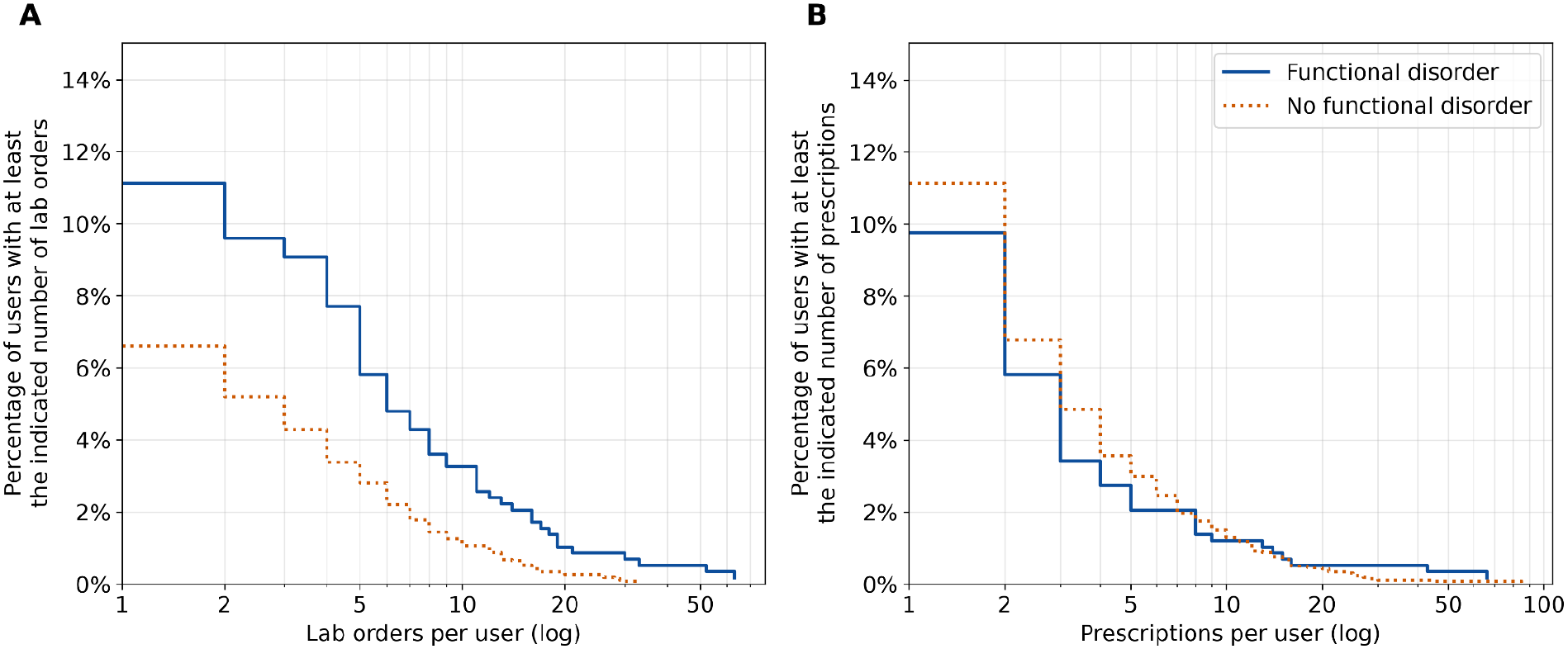
Lab orders and prescriptions for Lotus users with and without functional disorders. Panel A shows lab orders and Panel B shows prescriptions per user. Each curve shows the percentage of users with at least the number of lab orders or prescriptions indicated on the horizontal axis, which is plotted on a logarithmic scale; the value at 1 is the percentage of users with any lab order or prescription. The solid line shows users with a functional disorder, and the dotted line shows users without a functional disorder. Curves that lie higher or extend farther to the right indicate that a larger share of users had more lab orders or prescriptions.

### Care fragmentation

In all, 952 users (29.5%) had at least four prior visits with an identifiable provider; they averaged 8.8 visits across 4.8 providers. The most frequently seen provider accounted for a median of 40% of a user’s visits (UPC 0.40; IQR 0.29 to 0.54). 44.4% of users had high fragmentation indicated by rBBI (43.6% at 18 to 44 years, 45.9% at 45 to 64, and 43.7% at 65 or older; Table 2).

**Table 2.** Prior-year healthcare utilization and care fragmentation among Lotus users, by age group and functional disorder diagnosis.

|  |  |  |  |  | **rBBI category, %<sup>e</sup>** |  |  |
|---|---|---|---|---|---|---|---|
| **Group** | **n (%)<sup>a</sup>** | **Mean visits<sup>b</sup>** | **Mean providers<sup>c</sup>** | **Median UPC<sup>d</sup>** | **High (>0.85)** | **Medium (0.70–0.85)** | **Low (<0.70)** |
| **All users** | **952 (100.0)** | **8.83** | **4.78** | **0.40** | **44.4** | **29.7** | **25.8** |
| **Age, years** |  |  |  |  |  |  |  |
| 18–44 | 257 (27.0) | 7.84 | 4.42 | 0.43 | 43.6 | 24.5 | 31.9 |
| 45–64 | 329 (34.6) | 8.57 | 4.76 | 0.40 | 45.9 | 28.9 | 25.2 |
| 65+ | 366 (38.4) | 9.77 | 5.05 | 0.40 | 43.7 | 34.2 | 22.1 |
| **Functional disorder diagnosis** |  |  |  |  |  |  |  |
| Yes | 302 (31.7) | 10.75 | 5.75 | 0.36 | 53.6 | 24.8 | 21.5 |
| No | 650 (68.3) | 7.94 | 4.33 | 0.43 | 40.2 | 32.0 | 27.8 |

Abbreviations: rBBI, reversed Bice-Boxerman Index; UPC, Usual Provider of Care index.
<sup>a</sup> Percent of the analytic cohort (N = 952): Lotus users with ≥4 ambulatory visits in the 12 months before enrollment.
<sup>b</sup> Mean number of ambulatory visits per user in the 12 months before enrollment.
<sup>c</sup> Mean number of distinct clinicians seen per user over the same period.
<sup>d</sup> UPC is the proportion of a user's visits made to their most frequently seen provider (range 0–1); higher values indicate greater continuity of care.
<sup>e</sup> rBBI is 1 minus the Bice-Boxerman Continuity of Care Index, which reflects both the number of distinct providers and how evenly visits are spread across them (range 0–1); higher values indicate greater fragmentation.

### Functional disorders and care fragmentation

Users with a functional disorder were more likely to meet the four-visit threshold (51.7% vs. 24.6%; age- and sex-adjusted OR, 3.07; 95% CI, 2.52 to 3.72; P<0.001) and, among those assessed, more likely to have high fragmentation (53.6% vs. 40.2%; OR, 1.72; 95% CI, 1.30 to 2.29; P<0.001), an association that persisted after adjustment for age, sex, and visit count (adjusted OR, 1.41; 95% CI, 1.05 to 1.89; P=0.02). They made up 38.3% of the high-fragmentation group vs. 26.5% and 26.4% of the medium and low groups. Chronic conditions, by contrast, predicted inclusion in the analysis (adjusted OR, 3.77; 95% CI, 3.07 to 4.61) but not high fragmentation (adjusted OR, 1.12; 95% CI, 0.76 to 1.65; P=0.55).

## Discussion

Users of our care platform were older and predominantly male, with substantial chronic disease burden, functional disorder prevalence, and fragmented prior care. These observed differences from other users of digital-first care^12,13,34^ suggest that removing barriers of cost, scheduling, and gatekeeping may attract a unique population with distinct care needs and support the transferable lesson that routine characterization of deployed AI care populations against national benchmarks is critical to inform evaluation, safety monitoring, and quality improvement. Our study reports our approach to evaluating users of an AI health product and identifies three priorities for our platform and the broader development of LLM-enabled primary care.

First, the high chronic disease burden in our cohort calls for a platform designed to support monitoring, treatment adjustment, and coordination across conditions. Chronic disease management is a potentially high-value use of AI-led care, but must be considered a core component of the platform’s workload rather than an edge case. Early studies suggest AI-led chronic disease management is feasible, but evidence remains limited around long-term clinical effectiveness, cultural competency, and algorithmic bias.^35,36^

Second, functional disorders need dedicated workflows that address diagnostic uncertainty and guide appropriate testing and referral. These conditions are difficult to diagnose and manage, drive disproportionate acute and chronic care utilization,^37^ and have risen in incidence since COVID-19,^38–41^ even as primary care capacity has shrunk.^42^ On our platform, users with functional disorders generated disproportionately more escalations and laboratory orders, but not more prescriptions, the same pattern of uncertainty-driven testing described in outpatient settings. A platform that can assemble scattered records and synthesize long histories is well placed to help, but only if grounded in evidence-based practices^43^: validating explanations of symptoms, testing informed by prior workups, and stepped referral to psychological care when indicated. These safeguards are necessary to avoid amplifying low-yield testing.

Third, substantial care fragmentation highlights the need to coordinate care across providers and evaluate whether the platform improves continuity. Among assessed users, care was spread across nearly five providers, and the share with highly fragmented care (44%) exceeded estimates for Medicare beneficiaries (26%)^44^ and older Veterans (33%).^45^ Fragmentation was similar across age bands, including adults aged 18–44, a group rarely studied. Users with functional disorders had greater fragmentation even after adjustment for visit count, reinforcing the need for workflows that address both clinical management and care coordination. For the platform, this means routing escalations to a consistent clinical team and sharing summaries across providers. We are working to assess fragmentation at enrollment and over time as one indicator of how our platform influences care continuity through information sharing and integrated care coordination.

## Limitations

This cross-sectional study describes self-selected users of a single platform, limiting generalizability to other settings and populations. Comparisons with national estimates should be interpreted cautiously because diagnoses identified through ICD-10 codes may differ from self-reported conditions in surveys such as NHIS. Our literature-informed definition of functional disorders has not been formally validated. Fragmentation estimates apply only to users with at least four prior visits and may be affected by year-to-year variation in utilization and differences in care capture between linked records and comparator claims data. Finally, we did not evaluate clinical outcomes or whether the platform improves care.

## Conclusion

Users of our personalized agentic health platform with physicians-in-the-loop had high chronic and functional disease burdens and arrived with highly fragmented prior care. These findings highlight the need for chronic disease management, dedicated functional disorder workflows, and care coordination in LLM-based care platforms. To achieve the promise of safe, equitable, and high-value care, we believe AI-led platforms need to characterize their users early and against national benchmarks, across demographics, chronic and functional disease burden, and fragmentation. Capturing and reporting these profiles should inform safety thresholds, care pathways, and evaluation metrics to ensure these platforms are designed for the needs of the patients they actually serve.

## Data Availability

All data produced in the present study are available upon reasonable request to the authors.

