## Supplementary material for "Enrollees and Early Lessons from an Agentic Healthcare Platform with Physicians in the Loop": (Table S1), (Table S2)

**Table S1: Diagnosis codes used to identify NHIS chronic conditions**

| **NHIS Conditions**[1,2] | **NHIS Question Wording**[3,4] | **ICD-10-CM Code / Range**[5,6] |
|---|---|---|
| Hypertension | "Have you EVER been told by a doctor or other health professional that you had... hypertension, also called high blood pressure?" | I10<br>(related: I11–I16, I1A.0) |
| Coronary Heart Disease | "Have you EVER been told by a doctor or other health professional that you had... coronary heart disease?" | I25.1-, I25.5, I25.6, I25.7-, I25.8-, I25.9<br>Heart attack: I21.-, I22.-, I25.2 |
| Stroke | "Have you EVER been told by a doctor or other health professional that you had... a stroke?" | I60–I63, I69.-, Z86.73 |
| Diabetes | [Codebook label] "Ever had diabetes". Women are first told "Other than during pregnancy". | E08–E13 |
| Current Asthma | "Have you EVER been told by a doctor or other health professional that you had asthma?" then "Do you still have asthma?" | J45.-, J82.83 |
| COPD, Emphysema, or Chronic Bronchitis | "Have you EVER been told by a doctor or other health professional that you had... COPD, emphysema, or chronic bronchitis?" | J41–J44 |
| Arthritis (incl. RA, Gout, Lupus, Fibromyalgia) | "Have you EVER been told by a doctor or other health professional that you had... some form of arthritis, rheumatoid arthritis, gout, lupus, or fibromyalgia?"[7] | M05–M06, M08, M10, M1A, M15–M19, M32.1-, M32.8, M32.9, L93.0–L93.2, M45, L40.5-, M79.7 |
| Cancer / Malignancy | "Have you EVER been told by a doctor or other health professional that you had... cancer or a malignancy of any kind?" | C00–C96, Z85.- |
| Hepatitis | "Have you EVER been told by a doctor or other health professional that you had... hepatitis?"[8] | B15–B19, K70.1-, K71.3–K71.5-, K73.-, K75.4 |
| Weak or Failing Kidneys | "Have you EVER been told by a doctor or other health professional that you had... weak or failing kidneys?" Do not include kidney stones, bladder infections, or incontinence.[7] | N18.-, N19 |

**Note.** Codes with dashes (e.g. I25.1-) indicate that subcategories are included.

**Table S2: Definition of functional disorders, ICD-10 codes, and prevalence among Lotus users**

| Disorder Category | Functional Disorder Name | ICD-10-CM Code | References |
|---|---|---|---|
| Gastrointestinal | Irritable Bowel Syndrome | K58.0, K58.1, K58.2, K58.8, K58.9 | Burton 2020[9]; Rometsch 2024[10]; van der Meulen 2024[11] |
| Central Sensitivity & Pain | Fibromyalgia | M79.7 | Burton 2020[3]; Rometsch 2024[10]; van der Meulen 2024[11] |
| Neuroimmune & Systemic | Myalgic Encephalomyelitis / Chronic Fatigue Syndrome | G93.32, R53.82<br>Before Oct 2022: G93.3 | Burton 2020[11]; Rometsch 2024[10]; van der Meulen 2024[11] |
| Gastrointestinal | Functional Dyspepsia | K30 | Rometsch 2024[10] |
| Gastrointestinal | Functional Constipation | K59.04 | Rometsch 2024[10] |
| Gastrointestinal | Functional Diarrhea | K59.1 | Rometsch 2024[10] |
| Gastrointestinal | Cyclic Vomiting Syndrome | G43.A0, G43.A1, R11.15 | Kovacic & Li 2021[12] |
| Urological & Pelvic Pain | Interstitial Cystitis / Bladder Pain Syndrome | N30.10, N30.11 | Akiyama 2020[13]; Warren 2014[14] |
| Neurological & Functional | Functional Neurological Disorder / Conversion Disorder | F44.4, F44.5, F44.6, F44.7 | Espay 2018[10]; Hallett 2022[15]; Rometsch 2024[10] |
| Musculoskeletal & Craniofacial | Temporomandibular Disorder (pain-related TMD only) | M26.621, M26.622, M26.623, M26.629, M79.11 | Slade 2020[16]; Yakkaphan 2023[17] |
| Post-Acute Infection | Long COVID (Post-COVID Condition / Post-Acute Sequelae of SARS-CoV-2 Infection) | U09.9 | Davis 2023[18]; Ely 2024[19]; Goldenberg 2024[20] |
| Autonomic & Cardiovascular | Postural Orthostatic Tachycardia Syndrome | G90.A (from Oct 2022)<br>Before Oct 2022: G90.9 | Castells 2024[21]; Goldenberg 2024[20]; Kulin 2026[22] |

| | | | |
|---|---|---|---|
| Immune & Allergic | Mast Cell Activation Syndrome | D89.40, D89.41, D89.42, D89.43, D89.44, D89.49 | Castells 2024[21] |
| Central Sensitivity & Pain | Complex Regional Pain Syndrome, type I | G90.50, G90.511–G90.519, G90.521–G90.529, G90.59 | Littlejohn & Guymer 2019[23]; Popkirov 2019[24]; Treede 2019[25] |

**Note.** The 14 conditions in this table are grouped as functional disorders for analysis, but the cited literature does not classify all of them the same way. Nine are *supported*, meaning at least one cited source names the condition as functional or applies functional diagnostic criteria: irritable bowel syndrome, fibromyalgia, myalgic encephalomyelitis/chronic fatigue syndrome, functional dyspepsia, functional constipation, functional diarrhea, cyclic vomiting syndrome, interstitial cystitis/bladder pain syndrome, and functional neurological disorder. Five are *adjacent*: pain-related temporomandibular disorder, long COVID, postural orthostatic tachycardia syndrome, mast cell activation syndrome, and complex regional pain syndrome type I. This exploratory group of overlapping conditions shares symptoms and frequent co-occurrence with functional somatic disorders but are not clearly classifiable as functional.
